# Identifying Rare Germline Variants Associated with Metastatic Prostate Cancer Through an Extreme Phenotype Study

**DOI:** 10.1101/2025.04.28.25326584

**Authors:** Yen-Yi Lin, Hamideh Sharifi Noghabi, Stanislav Volik, Robert Bell, Funda Sar, Anne Haegert, Hee Chul Chung, Ladan Fazli, Htoo Zarni Oo, Mads Daugaard, Ming-Han Kuo, Sheng-Chieh Hsu, Eddie L. Imeda, Claudio Zanettini, Lucio Queiroz, Balthasar Schlotmann, Kazzem Gheybi, Colin Cooper, Zsofia Kote-Jarai, Rosalind Eeles, Pan Prostate Cancer Group (PPCG), Hsing-Jien Kung, Luigi Marchionni, Joachim Weischenfeldt, Keith D. Miller, Alan Rabinowitz, Yuzhuo Wang, Hai-Feng Zhang, Poul H. Sorensen, Mark S. Carey, Martin Gleave, Vanessa M. Hayes, William T. Gibson, Colin C. Collins

## Abstract

**Background:** Studies of germline variants in prostate cancer (PCa) have largely focused on their connections to cancer predisposition. However, an understanding of how heritable factors contribute to cancer progression and metastasis remain limited.

**Objective:** To identify low frequency to rare germline nonsynonymous variants associated with increased risk for metastatic PCa (mPCa), while providing functional validation.

**Design:** We assembled an extreme phenotype cohort (EPC) of 52 patients diagnosed with predominantly high-grade (Gleason Score (GS) ≥ 8) PCa and > 7 years of follow-up for which localized treatment naïve tumor tissues were available. In half of the cases, the tumor had metastasized to bone, providing an even distribution of bone mPCa and non mPCa cases. Tumor and matched distant benign DNA samples were exome sequenced and analyzed for germline variants with population-wide minor allelic frequencies ≤ 2%. Findings were validated using two independent PCa germline cohorts, including a closely matched Australian study biased to aggressive disease (n = 53) and Pan Prostate Cancer Group (PPCG, n = 976). Two mPCa-promoting candidate variants in *KDM6B* and *BRCA2* were engineered into cell lines and functionalized.

**Results:** Germline nonsynonymous rare variants (gnsRVs) identified in 25 DNA Damage Repair (DDR) genes were significantly enriched in the mPCa patients (p=4.57e-06). Conversely, the prevalence of synonymous variants at minor allele frequencies of ≤ 2% were similar between the mPCa and non mPCa patients. The predictive power of variants in 53 non-DDR genes was validated in the Australian cohort (p=0.028) and correlated with high-risk PCa in PPCG (p=0.03). *KDM6B* K973Q showed functional significance despite being annotated as benign in ClinVar, while *BRCA2* I1962T showed sensitivity to Olaparib. In total, six EPC variants related to DNA repair or epigenetics were found to alter enzymatic activity.

**Conclusions:** EPCs coupled with low frequency/rare variant analyses may advance understanding of interactions between the germline and tumor in PCa. We identified a series of germline variants that were enriched among mPCa patients. Moreover, we showed that one of these variants confers a metastatic phenotype. Our findings suggest that germline testing at diagnosis may improve treatment stratification in PCa.

**Patient summary:** The presence of specific genetic variants among men with PCa may elevate the risk of mPCa once PCa develops. Knowledge of the variant burden at time of diagnosis may enable accurate stratification of some patients for aggressive therapeutic interventions.

## 1. Introduction

Prostate cancer (PCa) affects roughly 1.4 million men worldwide, with its lethal form, metastatic (mPCa), causing significant morbidity and resulting in ∼ 397,000 annual deaths worldwide [1], this is expected to worsen significantly with an aging population [2]. Most deaths are a consequence of high-risk, aggressive disease. Patients with high-risk localized PCa have a 50-70% probability of progressing to metastasis after local therapy [3], but the basis of this aggressive phenotype remains incompletely understood [4]. Identifying the genetic factors underlying the conversion from localized to metastatic disease is crucial for improving patient stratification, prognosis, and treatment.

Recent advances in PCa genomics and molecular biology have shed light on the molecular mechanisms underpinning therapy resistance and metastasis, including alterations in the androgen receptor (AR) signaling pathway, DNA Damage repair (DDR) genes, and the activation of alternative oncogenic pathways [5, 6] and the tumor microenvironment [7–9]. Despite these advances, there remains much to learn about the interplay of germline factors contributing to the aggressive PCa phenotype. A deeper understanding of these genetic determinants is poised to unlock novel targeted therapies and more personalized treatment stratification for PCa patients.

Rare variant analyses aim to uncover the contribution of genetic variants to disease causality [10]. Typically, rare (Minor Allele Frequency(MAF) <1%) and a lesser extent low-frequency variants (1%-5%), have larger effect sizes than common variants (>5%), accounting for a substantial proportion of the disease heritability, depending on context [11]. Also, they may play unique roles in complex genetic disorders, allowing for hypothesis-free identification of disease-associated gene function [12]. Generally, a variant is rare either because it has been selected against during evolution, or it is under neutral selection [13, 14]. In a recent study, Burns et al., whole genome sequenced 850 germlines from PCa patients to identify rare germline variants predictive of time to biochemical recurrence (BCR), a surrogate for aggressive disease [15]. They found rare deleterious coding germline variants in “Hallmark” cancer gene sets consistently associated with time to BCR, including independent cohort validation.

This use of rare variant analysis in extreme phenotypic cohorts (EPCs) is a powerful approach for identifying variants that contribute to aggressive disease, with particular relevance to cancer studies such as PCa which displays considerable clinical, biological and genomic heterogeneity [16]. EPC preselection aims to enrich the study population for rare variant discovery, providing a surrogate for classical study power [17]. Furthermore, by reducing phenotypic heterogeneity, EPC studies can minimize the effect of confounding factors, such as polygenic background or environmental influences [18]. This technique reduces the “noise” and amplifies causative genetic signals, thereby enabling the use of smaller cohorts [19].

In this study, we leveraged the power of combining an EPC comprised of mPCa and non-mPCa with rare variant germline analyses. This allows a smaller cohort size for variants with a MAF ≤ 2%, which are referred to as rare varaints. The EPC includes 52 ethnically and age-matched (mean age 65 years) treatment-naïve patients who underwent surgery for high-grade disease with > 7 years follow-up with (n=26) or without (n=26) bone metastasis. Ultimately, the identification of functionally-relevant rare variants provides valuable insights into the biological mechanisms underlying metastasis, with benefits to improve screening and rationale for targeted interventions.

## 2. Patients and Methods

### 2.1. EPC selection

An extreme phenotype cohort of 52 localized and predominantly (73%) pathologic high-grade (GS ≥ 8) formalin-fixed and paraffin-embedded (FFPE) tumor specimens was assembled from treatment-naive patients with a minimum of 7 years of follow-up. Outcomes data allowed for two study arms including patients presenting either with (n=26) or without (n=26) post-surgical bone metatstisis. We note that due to the challenge of assembling such a cohort, nine tumors (five metastatic, four non-metastatic) pathologically scored as GS 4+3 were included. All were GS ≥8 based on diagnostic biopsies. Five additional non-metastatic GS 3+4 tumors were also included by the pathologist based on diagnostic GS ≥8, advanced pathological stage (T3a), > 50% positive biopsy cores or positive surgical margins. Careful review identified a single GS 3+4 lacking high risk feature in both arms. The cohort arms were further matched for positive margins 10 *versus* 13, posivitive lymph nodes 1 *versus* 2, average PSA 11.8 (4.1-31.26) *versus* 11.3 (3.6-61), average biopsy GS 8 versus 8.1, average pathological GS 8.1 versus 8.4, average positive biopsy cores 3.6 versus 3.8 (Supplementary Table 1). The patient ancestry information, determined by Somalier [20], corresponded with the predominantly Caucasian and East Asian demographics in Vancouver (32 Caucasians and 13 East Asians (further details in Table 1.))

**Table 1.** Ancestry of the Vancouver Prostate Centre Extreme Phenotype Cohort (EPC), Australian, and Pan Prostate Cancer Group (PPCG) cohorts.

| Ancestry | Cohort Size | Caucasian | African American | African | East Asian and Others | South Asian |
| --- | --- | --- | --- | --- | --- | --- |
| VPC EPC | 52 | 61.5% | 0.0% | 0.0% | 34.7% | 3.8% |
| Australian Cohort | 53 | 100.0% | 0.0% | 0.0% | 0.0% | 0.0% |
| PPCG Cohort | 976 | 94.2% | 0.0% | 2.8% | 1.8% | 1.2% |

### 2.2 DNA Sequencing

Both tumor and respective distant matched benign prostate H&E sections were selected from different FFPE tissue blocks and reviewed by a research pathologist (LF). The cancerous regions were selected based on the highest pathological presenting tumor content. For DNA extraction, two to four 5 μm sections from archived tissue blocks were collected in 1.5 mL microcentrifuge tubes. The genomic DNA from tumour and matched distant benign tissues was extracted using the Maxwell RSC DNA FFPE kit and with the Maxwell RSC instrument (Promega) according to the manufacturer’s instructions. The quality and quantity of the extracted DNA were assessed with TapeStation 42100 (Agilent Technologies) and measured using the Qubit ssDNA Assay Kit (ThermoFisher), respectively. NEBNext FFPE DNA Repair Mix was used to minimize FFPE-related artifacts. Whole-exome sequencing (WES) was performed using the KAPA HyperCap workflow and SeqCap EZ MedExome panel (Roche NimbleGen) and the libraries were sequenced on the Illumina NextSeq500 (PE150). The mean sequencing depths achieved were 89X and 57X for tumor and benign samples, respectively. The identical WES protocol was applied to freshly collected cells for all clones in 2.7.

### 2.3 Variant Calling and Annotation

Sequencing data were aligned to the GRCh38 Human Genome (Ensembl Release 92) using BWA 0.7.17. PCR duplicates marked by Picard 2.18.11 and all pairs with more than five mapped mismatches (3% error rates) removed. Strelka2 2.9.10 [21] was employed for variant calling, and a variant was considered germline if it was covered by at least 15 reads with the alternative allele frequency (AAF, defined as the ratio of reads with alternative allele over the total number of reads covering the base) ≥0.2 in both tumor and matching benign tissues. Rare variants (defined here as MAF ≤ 2%) in both WGS and WES datasets were derived from population-wide gnomAD v.2.1.1 data [22]. Our analysis focused on nonsynonymous protein-altering variants (missense, nonsense, splice-site) in GENCODE v27. The deleteriousness of all alternative allele variants was assessed using integral methods, namely fathmm-MKL [23] and CADD PHRED scores [22].

### 2.4 Copy Number Variation (CNV)

We used Bionano Nexus Copy Number to determine the copy number profile for each tumour using the following settings: minimum read depth for CNV calculation – 20, minimum mapping quality – 30, minimum base quality – 20, 10^-6^ significance threshold, at least 3 calls for calling copy number status, maximum 1Mb distance between calls. The copy number status for genes was called only when a CNV region completely overlapped a gene’s genomic location using Nexus Copy Number Query functionality. We also applied scarHRD [24] to calculate Homologous Recombination Deficiency (HRD) scores that are based on loss-of-heterozygosity (LOH), telomeric allelic imbalance, and large-scale state transitions. Higher HRD scores imply defective DNA Damage Response (DDR) and potential sensitivity to PARP inhibitors.

### 2.5 Statistical analysis

#### 2.5.1 Burden and Variance Test

We use PPCG cohort to test the associations of detected gnsRVs with various clinical phenotypes (see Results). For each clinical phenotype, we will first test the association of every single gnsRV using the Wald test [25]. Next, collapsed all gnsRVs in a gene and test the gene-level associations using the Burden test. Finally, we aggregated the scores of all gnsRVs in a combined test for each phenotype using the sequence kernel association test (SKAT). SKAT, or variance component test in general, is known to be robust in the presence of opposite association directions and in the presence of many non-causal variants [26].

#### 2.5.2 Bootstrapping

To evaluate the significance of gnsRV enrichments in metastatic *versus* non-metastatic tumors within our EPC, we employed a bootstrapping approach. The observed enrichments in the 25 genes were compared to the enrichments of gnsRVs in an equal number of randomly selected protein-coding genes. This process was iterated 10,000 times to generate robust empirical distributions for both metastatic and non-metastatic samples. Subsequently, we compared the resulting distributions of gnsRV enrichments in target genes to those in randomly selected protein-coding genes using the Wilcoxon rank-sum test with continuity correction. This statistical analysis allowed us to assess the significance of the observed enrichments in the target genes relative to the background distribution of gnsRVs in the genome, providing confidence in our findings. Analysis of this nature should be robust to any evolutionary selection pressure that is not related specifically to the phenotype under consideration (i.e. mPCa *versus* PCa).

### 2.6 Validation Cohorts

The Australian cohort is a closely matched alternative EPC high-risk resource (86.8% of patients with aggressive diseases), with available WGS germline data (buffy coats) from 53 genetically confirmed European ancestral PCa patients. It also includes 21 metastases (17 bone or/and distant visceral (1 treated), 4 nodal), 12 BCR, 3 PSA and local recurrence, 3 non-curative radical prostatectomies (RP), and 14 cases reported as no BCR. [27]. The PPCG cohort was comprised of WGS germline data from 976 PCa patients: including 200 with metastases out of 531 high-risk (54.6%), 305 intermediate, and 140 low-risk of metastasis.

### 2.7 CRISPR primer editing and isolation of KDM6B knock-in clones

KDM6B K973Q was engineered into the wildtype (WT) *KDM6B* gene in the PCa cell line LNCaP cells using CRISPR/Cas9 prime editing[28, 29]. LNCaP cells were cultured in RPMI-1640 medium supplemented with 10% FBS and 1% P/S at 37°C in a 5% CO₂ incubator. The cells were seeded into 6-cm dishes 18 hours before transfection to ensure 70-80% confluency by the next day. The KDM6B_2917C pegRNA was cloned into the piggyPrime vector pPBT-PE2-PuroTK-pegRNA_GG [29]. For transfection, 2.5 µg of plasmid DNA was mixed in a 5:1 ratio (pegRNA:hyPBase) and delivered using 7.5 µl of PolyJet (SignaGen), following the manufacturer’s protocol. After transfection, cells were selected with 2 µg/ml puromycin. The oligonucleotides used for pegRNA cloning and KDM6B sequencing are listed below. Single-cell clones were isolated, and the editing efficiency verified using Sanger sequencing. Successful editing was confirmed in two independent LNCaP clones (homozygote C3 and heterozygote C6 knock-ins) selected for further study. WES analysis of these clones revealed no additional edits in the LNCaP cells. The edited LNCaP cells were grown in media with charcoal-stripped serum.

### 2.8 Cell migration and invasion assays

WT LNCaP and CRISPR edited clones C3 and C6 were grown to 90% confluency on a 96-well ImageLock plate (Essen BioScience Cat# 4379) pre-coated with 50 μL of 50 μg/mL Poly-D-Lysine. A uniform wound was created in the cell monolayer using the 96-pin IncuCyte Wound Maker Tool. The cells were then gently washed twice with PBS to remove debris, and the medium was replaced with serum-starved medium to minimize cell proliferation. The plates were incubated for 48 hours, during which the progression of wound closure was monitored and captured using the IncuCyte Live-Cell Analysis System (Sartorius). To better measure the effects of cell migration and invasion, respectively, in the context of metastasis, the Transwell/ Boyden chamber migration and invasion assays were utilized. WT LNCaP and C3 and C6 cells were seeded onto the inserts and grown using the starvation media. The insert placed in a well containing a medium enriched with growth factors or chemoattractants (e.g., serum), and the cells incubated in a humidified incubator at 37°C to allow migration for 24 hours. After incubation, migrated cells on the lower side of inserts were fixed using methanol for analysis. Once fixed, the migrated cells (located on the outer side of insert) are stained using Crystal violet, a nuclei-specific histological dye, and quantified. The percentage of cells in each insert were measured or quantified using ImageJ.

### 2.9 PARPi sensitivity. To test if BRCA2 variant rs1060502377 (I1962T) attenuates BRCA2 activity in response to PARPi

the variant was engineered into HEK293T cells to compare its sensitivity to Olaparib treatment with DMSO controls on cell viability using the CCK-8 assay Selleckchem #B34304 [30]. WT and BRCA2-edited HEK293T cells were seeded in 96-well plates at 5,000 cells per well and treated with 100 µM Olaparib or an equivalent volume of 0.1% DMSO as a control. After 48 hours of treatment, 10 µL of CCK-8 reagent was added to each well, and the cells were incubated for an additional 2 hours. Cell viability was assessed by measuring absorbance at 450 nm using a microplate reader, with the treated cells’ absorbance compared to that of untreated control cells.

### 2.10 RNA sequencing and biological characterizations

RNA-Seq data were aligned to the GRCh38 release 90 reference transcriptome using STAR v2.6.0a [31]. Gene expression was quantified with HTSeq v0.11.2 [32], and differential expression was analyzed using DESeq2 v1.16.1 [33]. Genes with a fold change of ≥ 2 or ≤ 0.5 and an adjusted p-value less than 0.05 were considered significantly differentially expressed. These genes were then tested for enrichment with Hallmark gene sets, canonical pathways, and ontology gene sets from MSigDB [34].

## 3. Results

Focused on gnsRVs (gnomAD population frequency ≤ 2%) that alter protein sequences (missense, stop-gain, stop-loss, splice-site variants), we identified 10,854 gnsRVs present in the mPCa, yet absent in the non mPCa study arm. mPCa-specific gnsRVs were further assessed for potential pathogenicity using fathmm-MKL[23], identifying 6,080 variants with potential to impact protein function. Of these, 56 variants from 53 genes appeared among at least three mPCa patients at 11.5% penetrence.

Our germline variant calling relies on DNA extracted from FFPE histologically benign prostate tissue distant from the tumor site (usually in another prostate quadrant). This method poses two potential problems, the first being sequence artifact caused by the FFPE protocol, and the second being tumor admixture in the normal surrogate DNA. The fixing process in FFPE is known to cause deamination and other chemical modifications of DNA that require repair prior to sequencing and variant calling [35]. To determine the efficiency of repair matched flash frozen (FF) tissues from 7 patients (3 and 4 are from the metastatic and nonmetastatic groups respectively) were exome sequenced and variants called. 97.72% of the FFPE calls were observed in FF tissues indicating very efficient repair of artifacts. Moreover, most sequence artifacts occurred at sites AAF < 10% whereas our strict cut-off for variants is AAF > 20% thus eliminating most remaining artifacts. Next, we physically ruled out tumor admixture in the DNA isolated from distant benign prostate tissue using CNV analysis. First, we used random, non-matched benign DNA as a baseline to derive tumor CNV profiles. In every case, the resulting CNV profiles were identical to those obtained using matched benign DNA as a baseline. Next, we generated CNV profiles from benign samples, using other benign samples as the baseline. In all cases, these profiles were flat, while the profiles from random tumor samples showed significant variation. These results combined with our variant calling threshold cut-off of AAF > 20% rule out significant tumor admixture in our normal germline surrogate DNA.

To evaluate the validity of the pipeline and test our hypothesis, we first assessed for DDR genes [36, 37]. DDR gene variants can disrupt DNA homologous recombination (HR), leading to a BRCA1/2 genome structure [38] or BRCAness [39]. Alterations in DDR genes are also among the most common genetic events observed in metastatic PCa [40]. We selected 25 DDR genes most relevant for PCa and compared DDR gnsRVs distributions between the two arms of the cohort. We identified 71 gnsRVs (81 total 6 recurrent) exclusive to the metastatic arm and 21 gnsRVs exclusive to the nonmetastatic arm. The gnsRV burden of these 25 DDR genes is significantly higher in the metastatic arm compared to the nonmetastatic arm (p-value=4.57e-06), which is evidence of strong association with metastasis **(Fig. 1A&B)**. For scientific rigor the 5 non metastatic patients having only diagnostic GS and five random metastatic patients were deleted and the burden analysis repeated (p-value=5.9e-05) remaining highly significant. As a control variant set, we investigated germline synonymous rare variants (gsRVs) between the mPCa and non-mPCa arms. Not only was there no difference between the two arms (10 *versus* 13) but there were far fewer gsRVs than gnsRVs in each arm **(Fig. 1C&D)**. This was further confirmed through bootstrapping based on gnsRVs from randomly selected protein-coding genes where the Wilcoxon rank sum test p-value for the metastatic cohort being enriched with DDR gnsRVs is <2.2e-16 whereas the non-metastatic cohort showed no significance.

**Figure 1.**
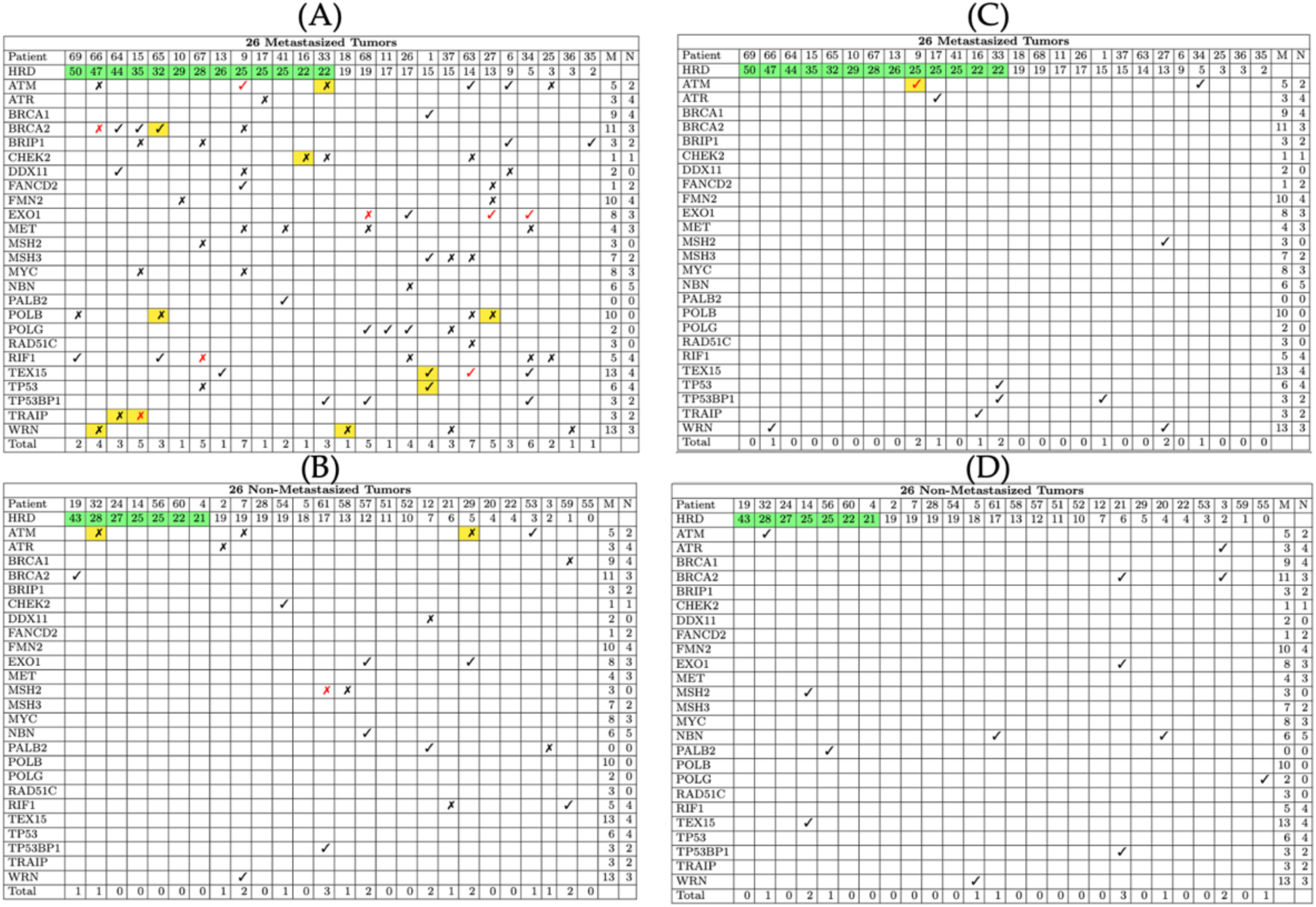
DDR gene gnsRVs and germline rare sysnonymous variants (gsRVs) in both groups of the EPC. Patterns of 102 DDR gnsRVs exclusive to either the metastatic **(A)** or non-metastatic **(B)** arm of the extreme phenotype cohort: 81 exclusive to mets **(A)** and 21 exclusive to non-mets **(B)**. Green cells: HRD scores ≥ 21. ✓: gnsRV. ✗: gnsRV predicted to be deleterious. ✓ or ✗ indicate compound gnsRVs. Yellow cells: allele frequency of the gnsRV in the matched tumor is at least 1.5-fold the allele frequency in benign suggesting positive selection. The column “M” and ”N” are the number of metastatic and nonmetastatic tumors with copy number aberrations detected at the gene locus. BRCA2 gnsRVs are 100% concordant with high HRD scores. The average HRD score is 21.58 vs. 14.62 (t-test p-value=0.04) in metastatic vs. nonmetastatic patients, respectively. There are fewer patients with HRD scores ≥21 (7 versus 13) in the non-metastatic cohort. The WRN gnsRV in patients 66 and 36 is known to impair helicase activity (PMID: 15489508). It is enriched 153-fold (2 out of 26 versus 0.0005 in gnomAD) over the general population. The POLB and CHEK2 variants are known to be deleterious(PMID: 23144635, 35127508). ***DDR gnsRV are enriched in metastatic vs non-metastatic tumors (p-value=4.57e-06)*** *using a Wilcoxon rank sum test) supporting positive selection for these gnsRVs in the metastatic arm.* No significant difference was detected for 11 synonymous RVs exclusive to mets in (C) and 13 exclusive to non-mets in (D).

A higher HRD score (≥21) implies defective DDR. To investigate if *BRCA2* variants associate with HRD, HRD scores were generated for all 52 tumors. The 26 metastatic tumors had a significantly higher combined HRD score (21.58 versus 14.62, t-test p-value=0.04), which is in line with PCa and monoallelic loss of *BRCA2* [41]. In addition, 13 of the 26 metastatic tumors had high HRD scores, compared to 7 in the nonmetastatic cohort. *BRCA2* gnsRV are 100% concordant with HRD scores ≥21 (including 1 non-metastatic tumor) suggesting impaired BRCA2 mediated DNA repair (**Fig. 1A&B**).

Using genome-wide interrogation we identified 56 gnsRVs in 53 genes **(Fig. 2)** that occurred exclusively among 3 or more patients in the metastatic arm suggesting that they are candidates for potentiating aggressive high-risk PCa. The variant burden within these 53 genes also showed a significant difference between the two patient groups (p-value = 2.98e-08), an observation that implies pre-existing germline variation within the 53 genes modified the risk of metastasis. If this is true, then at least some of these genes should be under a selection, positive or negative, in prostate tumors that develop during the lifetime of men who carry these gnsRVs. To test this, gene-level analysis was performed for a subset of the 53 genes in public PCa cohorts. Most tested genes were either mostly gained or lost suggesting biological selection in tumors (**Fig. 3A**). This provided evidence that these genes do undergo selection in PCa and collectively impact overall survival once PCa develops (p-value=1.88e-09.) **(Fig. 3A,B&C)**. Notable gnsRV include *POLB* rs3136797 (P242R, gnomAD MAF= 0.011) a DDR gene and a *KDM6B* rs61764072 (K973Q, gnomAD MAF= 0.009), a histone demythlase gene.

**Figure 2.**
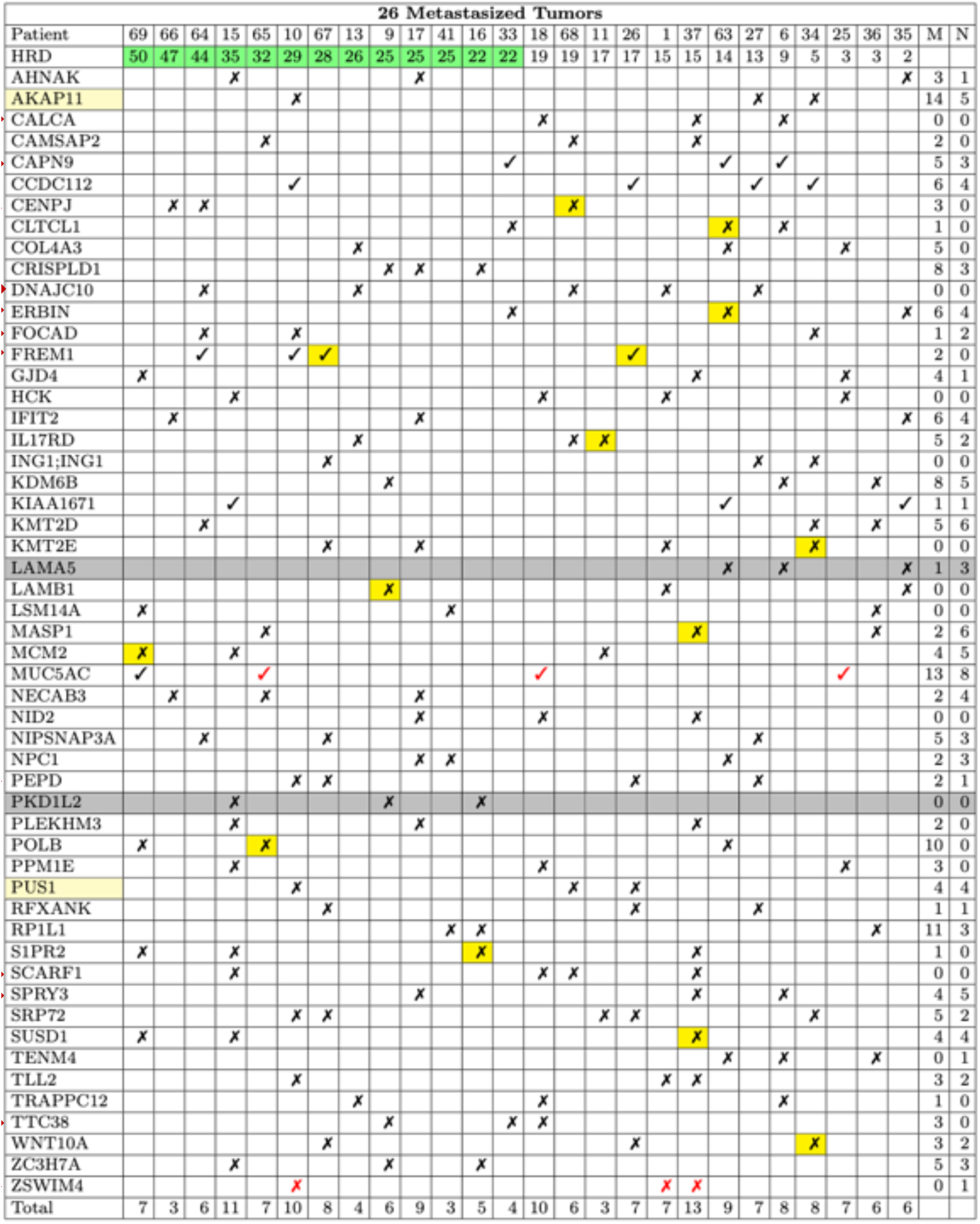
56 gnsRVs specific to and enriched in 26 localized metastatic tumors from 53 genes. Each gnsRV is recurrent in at least 3 (out of 26) tumors and is predicted or known to be deleterious. ✓ rare variant. ✗: predicted deleterious rare variant by fathmm-MKL. ✓ or ✗ indicate rare compound variants. Yellow cells: allele frequency of the gnsRV in the matched tumor is at least 1.5-fold the allele frequency in benign suggesting positive selection. The column “M” and ”N” are the number of metastatic and nonmetastatic tumors with copy number aberrations detected at the gene locus. Genes in yellow have their gnsRVs reported in COSMIC. The enrichment of the gnsRVs of in our VPC cohort ranges from > 10-fold to >200-fold over gnomAD population frequencies. The number of gnsRVs per patient is shown at the bottom. The average is ∼ eight. ***In the non-metastatic cohort, only LAMA5 and PKD1L2 had recurrent gnsRVs meeting our filtration requirement. These are shaded in gray***.

**Figure 3.**
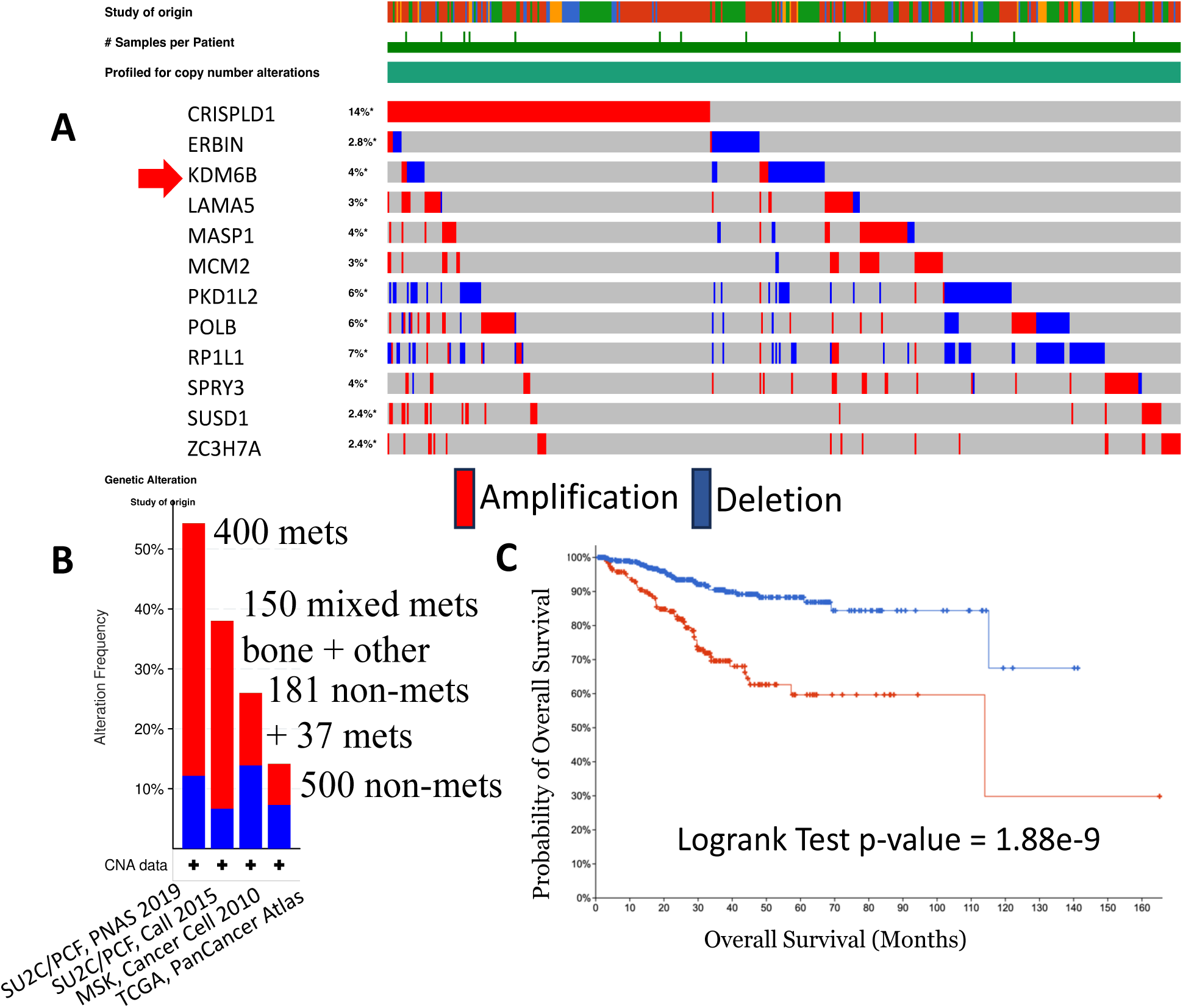
Genomic Analysis of Public PCa Cohorts for Genes with Recurrent gnsRVs in the EPC. Gene-level analysis was performed on 12 genes, selected from Figure 2, across 4 public prostate cancer (PCa) cohorts to determine if these genes are under selective pressure in PCa and collectively impact overall survival after disease onset. (A) A subset of genes with recurrent gnsRVs from Figure 2 was analyzed using cBioPortal for copy number variant analysis. Among 4 selected PCa cohorts, most genes are either predominately gained (e.g., CRISPLD1) or lost (e.g., PKD1L2) and thus appear to be under selection. (B) Copy number aberrations of 12 genes increase as tumors progress from adenocarcinoma to metastasis. This suggests that as tumors progress from relatively low risk of metastasis to a high risk of metastasis, these genes undergo positive selection. (C) Survival analysis of 12 genes among 4 PCa cohorts. Taken together, copy number aberrations at these genes predict a poor outcome with a Log-rank Test P-value of 1.88e-9. This suggests that these genes may play a role in the aggressive phenotype and impact patient outcomes.

Having developed the above Discovery Set of gnsRVs, we then sought a validation set within a similarly-enriched patient cohort. An independent and closely matched cohort of Australian PCa patients was interrogated for the presence of these 56 gnsRVs. The number of patients carrying at least one of these variants is higher than the expected carrier frequency as estimated using binomial distributions. The statistical significance for variants using our rare variant criteria (p=0.05 mets, 0.04 mets +BCR, and 0.25 non-mets) is consistently stronger in the metastatic phenotype group compared to the non-metastatic-phenotype group (**Table 3**). These results are concordant with the findings in our EPC, supporting that the recurrent variants identified in the EPC do associate with, and may contribute to the metastatic phenotype for high-risk patients.

**Table 2.** The allele frequency enrichment of germline nonsynonymous rare variants (gnsRVs) discovered from the VPC cohort in the independent PPCG cohort compared to healthy Caucasians (CAs) in gnomAD. We identified gnsRVs exclusive to metastatic patients in the EPC cohort and determined their population frequency within the independent PPCG cohort of 976 PCa germlines. We then compared these frequencies to those of healthy Caucasians (CA) in gnomAD. The average fold changes were 3.82, 4.42, and 5.86 for DDR gnsRVs, DDR gnsRVs predicted to be deleterious, and gnsRVs occurring at least three times, respectively. The average CA allele frequency closely matches the overall population allele frequencies reported in gnomAD. When considering only variants with ≥ 1.2-fold positive enrichment, we observed values of 8.25, 10.61, and 17.9. (t) Notes that the values of 5.86 and 17.9 in 3 recurrent gnsRVs are driven by one very rare gnsRV rs76761697 in the NIPSNAP3A gene that occurs in three PPCG germlines and in three metastatic patients from the EPC. If this gnsRV is removed, the values drop to 2 and 5-fold enrichment, respectively.

|  | Number<br><u>gnsRVs</u> | Total #<br><u>gnsRVs</u><br>(PPCG) | Average Allele<br>Frequency<br>(<u>gnomAD</u><br>Caucasian) | Average Fold Change In<br>Allele Frequency<br>(PPCG/ <u>gnomAD</u><br>Caucasians) | Average Fold Change In<br>Allele Frequency<br>(PPCG/<u>gnomAD</u> Caucasians)<br>**$\geq 1.2$ enrichment** |
| --- | --- | --- | --- | --- | --- |
| Variant-Centric: $\geq 3$ Recurrent<br><u>gnsRVs</u>* | 56 | 51 | 0.012 | 5.86<sup>t</sup><br>(0.16 ~ 199)<br>(21 variants enriched) | 17.9<sup>t</sup><br>(15 variants) |
| DDR<br><u>gnsRVs</u> | 81 | 38 | 0.004 | 3.82<br>(0.3 ~ 56.8)<br>(16 variants enriched) | 8.25<br>(13 variants) |
| DDR<br>Predicted<br>Deleterious | 45 | 21 | 0.004 | 4.42<br>(0.45 ~ 56.8)<br>(10 variants enriched) | 10.61<br>(7 variants) |

**Table 3.** Association of 56gnsRVs discovered from the EPC cohort in the independent Australian cohort of 53 Caucasian PCa germilnes. This independent CA cohort consisted of 21 metastatic cases (17 with bone and/or distant visceral metastases, and 4 with lymph node metastases), 12 with biochemical recurrence (BCR), and 14 without metastases (with clinical follow-up of more than 10 years). We determined the number of patients carrying at least one of these gnsRVs in each group and compared these frequencies to the expected carrier frequencies, estimated using binomial distributions. The enrichment is statistically significant for aggressive cancer groups, further supporting our hypothesis that the recurrent variants identified in the EPC cohort are associated with the progression of PCa to metastatic castration-resistant prostate cancer (mPCa).

|  | Cases with gnsRVs | Number of Patients | p-value |
| --- | --- | --- | --- |
| Met | 9 | 17 | 0.05063704 |
| Nodal | 2 | 4 | 0.3686052 |
| BCR | 6 | 12 | 0.3072268 |
| NonMet | 5 | 14 | 0.2493105 |
| Others | 3 | 6 | 0.2772393 |
| Met+BCR | 14 | 29 | 0.03995316 |
| Met+Nodal+BCR | 16 | 33 | 0.02779074 |

Finally, as a second Replication Set we analyzed germline WGS data from 976 PCa patients from the PPCG consortium for the presence of gnsRVs that include 81 gnsRVs in 25 DDR genes (Fig. 1A), 56 gnsRVs exclusive to and recurrent to our metastatic arm, and 343 gnsRVs from 206 genes with gnsRVs exclusive to (1-2 per germline) the metastatic patients, resulting in 476 unique gnsRVs. 356 of those were detected as germline variants in the PPCG cohort. We found at least 3X enrichment in PPCG relative to gnomAD Caucasian population frequency. We further tested gene-level associations with clinical-pathologic features in the PPCG cohort using burden tests. Significant associations with younger age at radical prostatectomy (*FOCAD*, *CHEK2*), PSA at RP (*ACSF3, SIVA1, HPGDS*), metastasis (*SPATA9*), and high risk (*ZSWIM4, DNAJC10*) were identified for the genes that contain selected gnsRVs. The combined score of all gnsRVs using SKAT test shows a significant association with high-risk disease in PPCG (p-value < 0.03) **(Table 4)**. This suggests that gnsRVs specific to the metastatic arm in EPC also correlate to aggressive PCa in the PPCG cohort.

**Table 4.** Representative genes containing gnsRVs that associated (nominal p-values) with phenotypes in the PPCG cohort. The FOCAD and CHEK2 gnsRV are linked to multiple cancers and may be associated with early-onset cancers. SIVA1 is an E3 ubiquitin ligase that regulates cell cycle progression, proliferation, and apoptosis in an ARF/p53-dependent manner. ACSF3 (PSA at RP, high-risk, and metastasis) is involved in fatty acids metabolism, and the gnsRV is likely pathogenic. SPATA9 associates with multiple clinical-pathologic features in the PPCG cohort, including metastasis. DNAJC10 is involved in misfolded protein degradation and is associated with poor survival in breast cancer and germline variants associated with aggressive PCa. *FOCAD, SIVA1, and DNAJC10 have higher minor allele frequency in African American populations than those of CAs* [62].

| Phenotype | Gene | P-value | Caucasian<br>MAF in <u>gnomAD</u> | African American<br>MAF in <u>gnomAD</u> |
| --- | --- | --- | --- | --- |
| Age at RP | *FOCAD* | .0000001 | **.0001** | **.0007** |
|  | *CHEK2* | .00001 | .005 | .00008 |
| PSA at RP | *SIVA1* | 10e-80 | **.00009** | **.0328** |
|  | *ACSF3* | 10e-80 | .001 | .0005 |
| Metastasis | *SPATA9*<sup>1</sup> | .005 | .0157 | .0022 |
| High-Risk | *ZSWIMJ*<sup>2</sup> | .001 | .0257 | .004 |
|  | *DNAJC10* | .001 | **.005** | **.027** |
| High-Risk | Aggregated <u>gnsRVs</u> | .03 | N/A | N/A |

We also highlighted the functional and clinical significance of gnsRVs identified in our study to demonstrate the effectiveness of our strategy. The *KDM6B* gnsRV rs61764072 (K973Q, gnomAD MAF=0.009) was selected to test whether variants annotated as “benign” in ClinVar [42] could still alter gene activity. KDM6B is a histone demethylase involved in epigenetic regulation. It targets and demethylates lysine 27 on histone H3 (H3K27me3), activating gene expression [43–45]. KDM6B is also known to play a role in PCa [46]. Given its involvement in eoigentic regulation, we hypothesized that the selected variant might impact global gene expression, making its effect easier to detect. The *KDM6BgnsRV* rs61764072 (K973Q) occurs in 11.5% of germlines of the EPC metastatic arm alone. While this variant lies outside of the catalytic JMJD domain and thus may not directly affectenzyme activity, it could alter interactions with other proteins to influence chromatin binding sites and gene expression patterns(**Fig. 4A**). AlphaFold modeling [47] suggests that rs61764072 is located on the protein surface, potentially functioning as a rotamer [48] (**Fig. 4B**). Analyses utilizing several predictive algorithms suggest that rs61764072 may modify KDM6B ability to reprogram global gene expression. To test that, we engineered the *KDM6B* K973Q variant into WT LNCaP cells using CRISPR/Cas9 prime editing [28, 29]. Successful editing was verified by WES, and two independent LNCaP clones (homozygote C3 and heterozygote C6) knock-ins were selected for further analysis. WES confirmed that rs61764072/ K973Q is the only variant present in the edited *KDM6B* gene and that no additional recurrent edits were introduced outside of *KDM6B*. The KDM6B protein abundance in LNCaP-C3 and LNCaP-C6 is comparable to that of the parental clone (**Fig 4C**).

**Figure 4.**
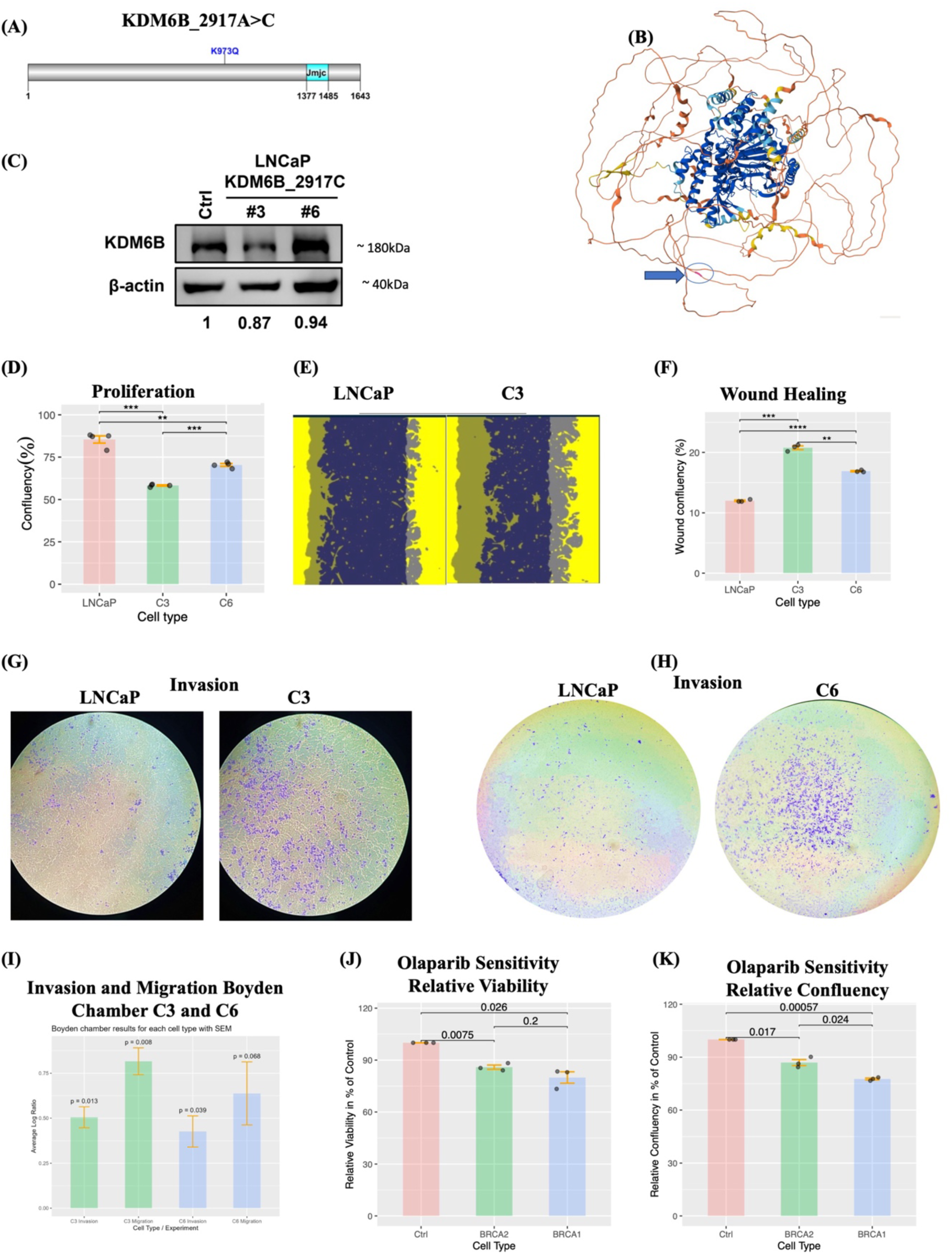
Functional assays and sequencing analysis of 2 gnsRVs discovered in the metastatic arm of EPC: KDM6B gnsRV rs61764072 (K973Q) in LNCaP and BRCA2 gnsRV rs1060502377(I1962T) in HEK293T. We edited two gnsRVs discovered in the EPC metastatic arm for functionalization. (A) KDM6B protein structure and location of the target gnsRV. KDM6B is a histone demethylase that demethylates lysine 27 on histone H3 (H3K27me3) thereby activating gene expression. The KDM6B gnsRV rs61764072 (K973Q) occurs in 11.5% of germlines of the extreme phenotype cohort metastatic arm (Fig. 2) and has a minor allele frequency approximately two-fold higher in the PPCG validation cohort compared to gnomAD. We hypothesized that KDM6B (K973Q) might impact global gene expression, making an effect straightforward to detect. rs61764072 (K973Q) lies outside the catalytic jmjd domain and may not directly impact enzyme activity, but it could influence interactions with other proteins, thereby affecting chromatin binding sites and gene expression patterns.(B) Despite its benign status in ClinVar, modeling using AlphaFold suggests an extrinsic positioning of rs61764072 (K973Q) on the protein surface, and several prediction algorithms predict it to be deleterious. To test this, we edited the K973Q variant into the KDM6B locus in wildtype LNCaP cells using CRISPR/Cas9 prime editing. (C) Two independent LNCaP clones, homozygote C3 and heterozygote C6 knock-ins, were selected for further study (successful editing in both was confirmed by whole exome sequencing.) The expression levels of KDM6B of LNCaP-C3 and LNCaP-C6 are comparable to that of the parental clone. (D) The growth rate of homozygous C3 is lower than that of heterozygous C6, with C6 growing slower than the WT. (E) Scratch assays reveal a mild migratory phenotype in edited LNCaP cells compared to WT. (G) Quantification shows C3 is more migratory than C6. (G,H,I) Boyden chamber assays reveal approximately a 4-fold increase in migration and invasion compared to WT LNCaP cells and again clone C3 (homozygous) displays stronger phenotypes than clone C6 (heterozygous) suggesting a dosage effect. (J,K) The BRCA2 gnsRV rs1060502377 (I1962T) (gnomAD MAF: 10e-7) confers sensitivity to PARP inhibitors, despite being classified as of unknown significance in ClinVar.

Proliferation assays revealed that clone C6 (heterozygous) grew faster that clone C3 (homozygous) (**Fig. 4D**). Scratch assays and Boyden chamber further demonstrated that the *KDM6B* K973Q confers both migratory and invasive phenotypes to LNCaP cells (**Fig. 4E-I**), with clone C3 exhibiting greater migration and invasion than C6. Remarkably, all assays showed a gene dosage effect, highlighting *KDM6B* K973Q-specific functional relevance. These findings provide evidence that *KDM6B* K973Q is functionally relevant, supporting our hypothesis that gnsRVs classified as benign or a VUS may, in a context-dependent manner, potentiate a metastatic phenotype in extant tumours.

A second rare variant (gnomAD MAF=0.00001), *BRCA2* rs1060502377 (I1962T), from the metastatic arm of the cohort, was assessed for attenuated activity in response to PARP inhibition. We showed that *BRCA2*-I1962T-edited HEK293T cells to be more sensitive to Olaparib compared to WT HEK293T and the positive control *BRCA1*-edited HEK293T cells (**Fig. 4J and K**). This suggests that *BRCA2*-I1962T may rely more heavily on PARP-mediated DNA repair mechanisms, making the patient carrier a promising candidate for PARP inhibitor treatment.

## 4. Discussion

While germline studies of common varaints have been instrumental in identifying genetic variants associated with increased susceptibility to PCa [49, 50], with the development of polygenic risk scores [51, 52], rare pathogenic variant identification is leading to an uptake in PCa germline testing [53, 54]. In contrast, the impact of germline variants on tumor phenotype have been largely overlooked - although this is changing [55–57]. Here, using our unique EPC high-risk metastatic *versus* non metastatic PCa study design, we interrogate for rare potentially pathogenic variants. Our study overcomes notable challenges associated with such studies, including the requirement for significant and lengthy clinical follow-up (over 7 years), the ability to generate accurate sequence data from FFPE specimens, and the use of matched distant benign tissue as a surrogate source of germline DNA. While our results clearly demonstrate that the use of pathologist-reviewed distant benign prostate tissue from FFPE blocks imposes no significant limitation to our study, our approach for technical validation shows the reliability of our FFPE sequenced data.

To test the hypothesis that combining an EPC study with a rare variant analysis can identify variants that impact the metastatic phenotype of a tumor, we assessed a panel of 25 DDR genes in each arm of the study for nonsynonymous and synonymous variants. This revealed an asymmetric distribution of nonsynonymous variants between the two arms (p-value=4.57e-06) **(Fig.1A&B)**, whereas no difference was detectable for synonymous variants. Bootstrapping indicates this frequency difference was unlikely to have occurred by chance (p-value <2.2e-16).

The analysis of variants identified in our EPC across independent cohorts further strengthens the link between our findings and PCa metastasis. A strict unbiased whole genome analysis was performed identifying 56 variants in 53 genes that are exclusive to and enriched in the metastatic arms. A subset of these genes is preferentially gained or lost in public PCa cohorts and predict for poor outcome, suggesting biological selection of these genes in tumors **(Fig.3A)**. These variants were first validated in a closely-matched Australian PCa cohort, where the number of patients carrying at least one of these coding variants exceeded the expected carrier frequency, as estimated using binomial distributions. The statistical significance for gnsRV carrier number was notably stronger in the mPCa phenotype group compared to the non mPCa group [p=0.05 mets, 0.04 mets +BCR and 0.25 non-mets] **(Table 3)**. We also analyzed the PPCG cohort of 967 PCa germlines including 200 mPCa and found significant enrichment for DDR variants, particularly those predicted to be pathogenic, as well as for some non-DDR genes. An association with metastasis was also identified (p=0.03). Rare variants in individual genes associated with age at RP (*FOCAD* and *CHEK2*), PSA at RP (*SIVA1* and *ACSF3*), metastasis (*SPATA9*), High Risk (*ZSWIM4* and *DNAJC10*). Three of these *FOCAD*, *SIVA1*, and *DNACJ10* had substantially higher population frequencies in the African American (AA) population. These investigations support our assertion that the recurrent germline variants identified in the mPCa EPC are linked to the tumor phenotype associated with the progression of PCa to mPCa.

The results of the gnsRV analysis in both our EPC and two independent cohorts suggests that the clinical and functional significance of variants of unknown significance (VUS) or benign gnsRVs are underestimated, and that the aggregate contributions of these gnsRVs interact with somatic mutations to influence outcome once an individual develops a PCa tumor. This combined low frequency/rare variant and extreme phenotype study identified variants in four DDR genes. These are *CHEK2* I157T [rs17879961], *POLB* P242R [rs3136797], *POLB* R137Q, and *WRN* R834C [rs3087425], all shown to alter enzymatic activity and/or impair DNA repair [58, 59]. The *POLB* variants are reported to induce genome instability, while the activity of the *CHEK2* variant is altered and segregates with familial nonmedullary thyroid cancer [60, 61]. In the PPCG cohort *CHEK2* I157T was associated with younger age at diagnosis (p-value=1.0e-06). We functionalized two variants annotated as benign and VUS in ClinVar in *KDM6B* and *BRCA2*, respectively [61]. Engineering the *KDM6B* K973Q into LNCaP dramatically altered global gene expression leading to a migratory and invasive phenotype in a dosage-dependent manner. Edited *BRCA2* I1962T [rs1060502377] in HEK293T conferred PARPi sensitivity on HEK293T. Significantly, PARPi sensitivity was intermediate between the negative and positive controls suggesting attenuated BRCA2 activity. Further studies are required to appreciate the detailed mechanism whereby rs61764072 and rs1060502377 modify tumor phenotypes.

## 5. Conclusion

The results of sequence analysis and functionalization demonstrate that our strategy, which combines EPC and low frequency/rare variant analyses, has the potential to uncover clinically and functionally significant variants related to PCa metastasis, even those annotated as benign. In aggregate low frequency and rare variants may be the equivalent of “death by a thousand cuts”. A potential shortcoming of this study is the small EPC size, however the results make a compelling case for much larger EPC studies aimed at delineating the contribution of the germline to shaping prostate tumor phenotypes.

## Data Availability

The raw data generated in the present study are currently under review in the SRA and will be updated upon approval. They are available upon reasonable request to the authors before public release.

## Ackonwledgement

C.S.C would like to thank those men with prostate cancer and the subjects who have donated their time and samples for this study. Part of the research presented in this paper was carried out on the High Performance Computing Cluster supported by the Research and Specialist Computing Support service at the University of East Anglia. We acknowledge support from Cancer Research UK C5047/A29626, C5047/A22530, C309/A11566, C368/A6743, A368/A7990, and C14303/A17197; We acknowledge support from Prostate Cancer UK (MA-TIA23-002, MA-ETNA19-003, TLD-CAF22-011), the Dallaglio Foundation; and Prostate Cancer Research. We also acknowledge support from the Bob Champion Cancer Research Trust, Movember, Big C Cancer Charity, the Masonic Charitable Foundation, the King Family, the Stephen Hargrave Foundation, Alan Boswell Group, The Bob Willis Fund and the Bedford Memorial Trust. R.A.E thank those men with prostate cancer and the subjects who have donated their time and their samples to the consortium. We also acknowledge support of the research staff in the groups who so carefully curated the samples and the follow-up data. We acknowledge support from Cancer Research UK C5047/A14835/A22530/ A17528, C309/A11566, C368/A6743, A368/A7990, C14303/A17197, Prostate Cancer UK (MA-TIA23-002, the Dallaglio Foundation (CR-UK Prostate Cancer ICGC Project and Pan Prostate Cancer Group), PC-UK/Movember, the NIHR support to The Biomedical Research Centre at The Institute of Cancer Research and The Royal Marsden NHS Foundation Trust.Genomic sequencing and interrogation of Southern African Prostate Cancer Study (SAPCS) data was supported by the Ancestry and Health Genomics Laboratory at the University of Sydney through National Health and Medical Research Council (NHMRC) of Australia funding, including a Project Grant (APP1165762) and Ideas Grants (APP2001098, APP2010551), and U.S.A. Congressionally Directed Medical Research Programs (CDMRP) Prostate Cancer Research Program (PCRP) funding, including an Idea Development Award (PC200390, TARGET Africa) and HEROIC Consortium Award (PC210168 and PC230673, HEROIC PCaPH Africa1K). Further support for SAPCS analytical costs was provided by the U.S.A. National Institute of Health (NIH) National Cancer Institute (NCI) Award (1R01CA285772-01) and a U.S.A. Prostate Cancer Foundation (PCF) Challenge Award (2023CHAL4150). V. M. H was further supported by the Petre Foundation via the University of Sydney Foundation (Australia). J. W was supported by the Danish Cancer Society #R147-A9843, Danish Cancer Society #R374-A22518, Danish Council for Independent Research #8020-00282, Danish Council for Independent Research #3101-00177A, Novo Nordisk Foundation #NNF200C0060141, Sygeforsikringen Danmark # 2022-0198. C.S.C, D.C.W, R.A.E, D.B and Z.K-J made a substantial contribution to the organisation and conduct of the study and critiqued the output for important intellectual content. H.J.K acknowledges the support of MOHW 114-TDU-B222-134002, NSTC-114-2634-039-001 and 114-2326-B-038-001 W.T.G gratefully acknowledges funding from the BC Children’s Hospital Research Institute IGAP award. C.C.C. acknowledges the support of Canadian Institue of Health Research PJT-175238 and PJT-153073, and the support of MITACS through the MITACS Accelerate program IT19627, in partnership with PHEMI Systems Corporation and the University of British Columbia.

## Abbreviations

AR: androgen receptor
BCR: biochemical recurrence
CNV: copy number variation
DDR: DNA damage repair
EPC: extreme phenotype cohort
FF: flash frozen
FFPE: formalin-fixed and paraffin-embedded
GS: Gleason Score
HR: homologous recombination
HRD: homologous recombination deficiency
LOH: loss-of-heterozygosity
MAF: minor Allele Frequency
PCa: prostate cancer
PSA: prostate-Specific Antigen
WES: whole-exome sequencing
WT: wildtype
gnsRV: germline nonsynonymous rare variants
mPCa: metastatic PCa

